# Patient and Clinician Perceptions of a “People-Like-Me” Tool for Personalized Rehabilitation after Total Knee Arthroplasty: A Qualitative Interview Study

**DOI:** 10.1101/2023.10.23.23297404

**Authors:** Laura Churchill, Jeremy Graber, Meredith Mealer, Charles A. Thigpen, Dan D. Matlock, Michael J. Bade, Jennifer E. Stevens-Lapsley

**Affiliations:** Eastern Colorado VA Health Care System, Geriatric Research Education and Clinical Center (GRECC), Aurora, CO, USA; Physical Therapy Program, Department of Physical Medicine and Rehabilitation, University of Colorado, Aurora, CO, USA; Department of Physical Medicine and Rehabilitation, School of Medicine, University of Colorado, Aurora, CO, USA; Mental Illness Research Education and Clinical Center, VA Eastern Colorado Healthcare System, Aurora, CO, USA; ATI Physical Therapy, Greenville, South Carolina

**Keywords:** total knee arthroplasty, rehabilitation, clinical decision support, personalized, prediction, expectations, interviews

## Abstract

**Objective:** We developed a decision support tool to help physical therapists (PTs) address the varied expectations and recoveries of the total knee arthroplasty (TKA) population. The purpose of this study was to explore patients’ and clinicians’ perceptions and experiences with the tool during rehabilitation after TKA.

**Methods:** We piloted the tool in 2 outpatient physical therapy clinics. We conducted in-depth semi-structured interviews with 1) patients who underwent TKA and were exposed to the tool during rehabilitation and 2) clinicians who used the tool with patients after TKA. Two members of the research team coded the interview data using a descriptive content analysis.

**Results:** 16 patients and 10 clinicians were interviewed. We identified 4 common themes: 1) Expectations: most patients and clinicians felt the tool provided patients with valuable feedback for managing recovery expectations; 2) Motivation: patients and clinicians felt the tool motivated patients to participate in rehabilitation by providing positive reinforcement and/or a form of competition; 3) Influence on practice: some patients and clinicians indicated that the tool helped guide treatment decisions or provided opportunities for patient education, but most felt it did not influence clinical decision making; and 4) Clarity and comprehension: the majority of patients understood the tool’s “take-home” message, however, some patients and clinicians felt the use of percentiles, line graphs, and medical jargon decreased patients’ clarity and comprehension of the tool.

**Conclusions:** Overall, participants reported that the tool helped to shape patients’ expectations for postoperative recovery and increase patient motivation to participate in rehabilitation. Participants had mixed perceptions on how the tool influenced clinical care. Finally, participants identified some limitations in patient comprehension of the tool, which will inform future revisions to the tool to accommodate varying levels of health literacy.

## Introduction

Total knee arthroplasty (TKA) improves pain and disability for many patients with knee osteoarthritis,^1^ but up to 20% of patients feel dissatisfied after surgery.^2^ Unmet expectations are a leading cause of dissatisfaction after TKA,^3–5^ which suggests TKA care should focus on (a) providing patients with realistic expectations about recovery and (b) helping patients meet their goals after surgery. However, addressing expectations after TKA can be challenging, because patients vary considerably in their postoperative goals and course of recovery. ^6–8^ Many patients also report feeling uncertain or under-informed about the recovery process after TKA,^9^ which can increase anxiety during an already stressful time.

Physical therapists (PTs) routinely help set patient expectations after TKA. PTs estimate patients’ recovery prognosis and use measures, like knee range of motion (ROM), to monitor recovery and set rehabilitation goals. PTs largely rely on clinical experience and generic evidence (e.g., surgical protocols, population recovery benchmarks) to estimate and monitor recovery. Unfortunately, this reliance on generic evidence may lead to unrealistic expectations and/or suboptimal treatment plans for some patients^10^—especially those whose recovery differs from the population average—and may contribute to post-TKA dissatisfaction.

We developed and validated a “people-like-me” tool to help PTs address the varied expectations and recoveries of the TKA population.^7,11,12^ The “people-like-me” approach is a framework that promotes person-centered care by “using historical outcomes data from similar (past) patients as a template of what to expect for a new patient”.^13,14^ Essentially, the “people-like-me” tool provided individualized projections of TKA recovery using the recovery data from similar historical patients. We envisioned PTs could use this tool to (1) help patients set realistic expectations about recovery, (2) monitor patients’ recovery compared to similar patients to stimulate personalized treatment and shared decision making, and (3) provide patients with context and feedback about their recovery to reduce postoperative anxiety and uncertainty.^15^

We piloted the “people like me” tool in two outpatient physical therapy clinics to explore its use in real-world rehabilitation practice. The purpose of this study was to explore patients’ and clinicians’ perceptions and experiences interacting with the tool during rehabilitation after TKA.

## Methods

### Description of tool

The tool consisted of a web-based interface that allowed clinicians to predict and monitor patient recovery using a “people-like-me” approach. Essentially, the tool used an algorithm to identify a subset of patients from a large historical database who were similar to a new patient. Then, it used the actual recovery data from these similar patients to predict the new patient’s recovery. These predictions were presented to patients and clinicians as “people-like-me” reference charts, which are conceptually similar to childhood growth charts. These “people-like-me” reference charts (1) displayed the patient’s projected recovery (including the uncertainty around this projection) and (2) compared the patient’s observed recovery against this projection in terms of percentiles. To make these reference charts more interpretable, the tool also provided text-based interpretations of the patient’s recovery (see Figure 1). The tool generated reference charts for commonly collected outcomes after TKA including knee flexion and extension range of motion (ROM), Timed Up and Go (TUG),^16^ and the Western Ontario & McMaster Universities Arthritis Index (WOMAC) pain subscale.^17^ The tool did not provide clinicians with specific treatment recommendations. Instead, clinicians were encouraged to use their clinical judgment to determine how the information should be applied. We have created a pared-down, open access version of the tool for knee flexion and TUG recovery that can be viewed at https://cu-restore.shinyapps.io/knee_recovery_v1/.

**Figure 1 -.**
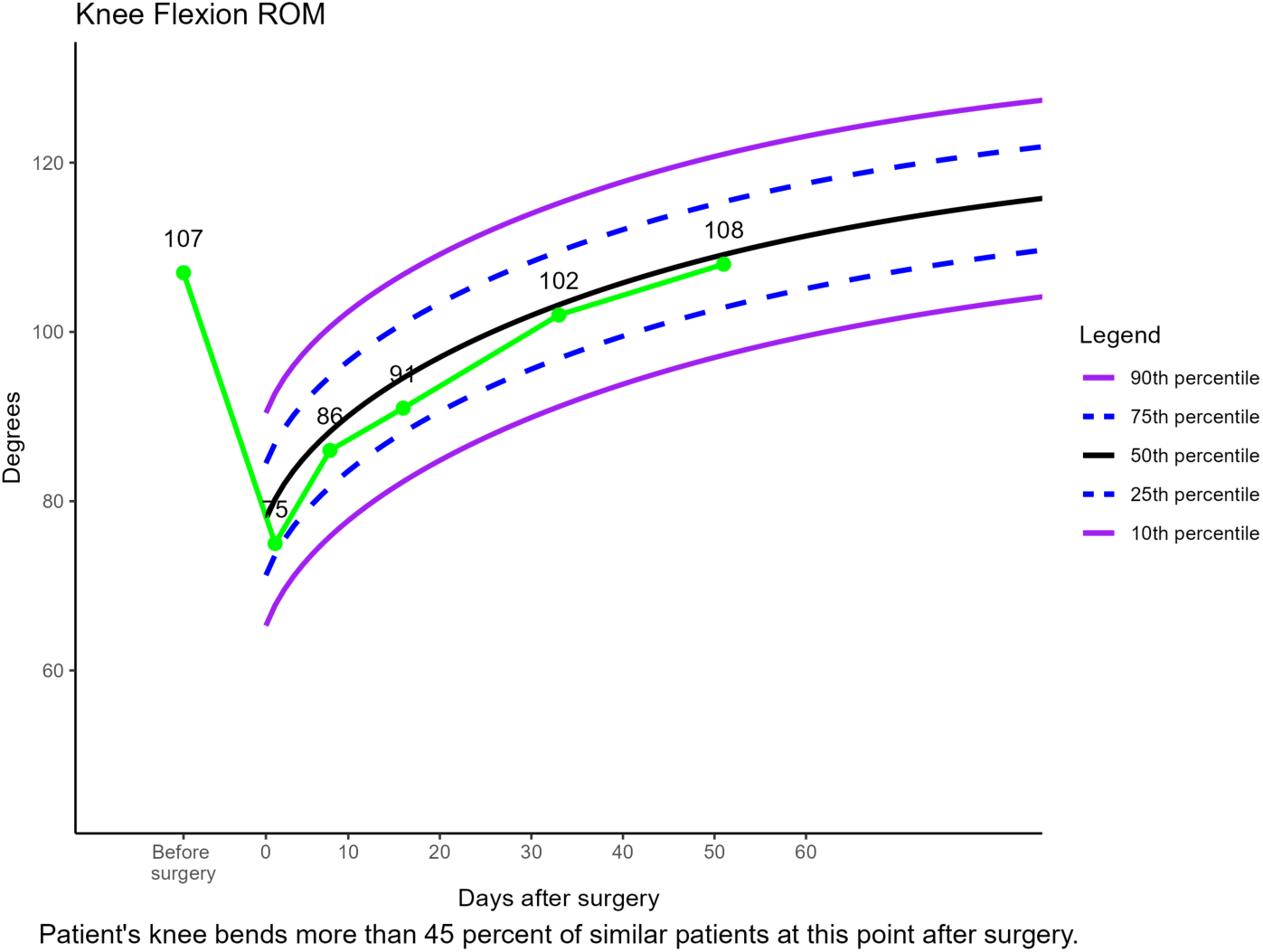
Example of the clinical decision support tool’s output

### Study design and setting

Two outpatient physical therapy clinics piloted the tool from September 2020 – December 2022. Both clinics are in the Greenville, South Carolina, USA metro area and belong to the same system of outpatient physical therapy clinics. The clinics have been collaborating with the research team since 2013 and contributed most of the data used to train the tool’s predictive algorithms. All permanent, full-time physical therapy clinicians (physical therapists and physical therapist assistants) were encouraged to use the tool with all patients after TKA. We have previously described the training and strategies used to support the tool’s implementation in a separate manuscript.^18^

Two members of the research team (LC and JG) conducted in-depth semi-structured interviews with two groups of participants: 1) patients who underwent TKA and who were exposed to the tool by their clinician(s) during rehabilitation and 2) clinicians who used the tool with patients after TKA. During this study LC was a physical therapist and postdoctoral researcher with experience in qualitative analysis. JG was a physical therapist and PhD candidate who was involved in the development of the tool and had previous exposure to clinician participants. All study procedures were approved by the Colorado Multiple Institute Review Board (COMIRB # 18-1246).

### Participant recruitment

We recruited a convenience sample of patients and clinicians (physical therapists and physical therapist assistants) from the pilot clinics. Patients were eligible to participate if they had been exposed to the tool (n=167) during their treatment and had been discharged from rehabilitation. Clinicians were eligible if they had used the tool with patients (n=16). We contacted patients (telephone and/or text) and clinicians (e-mail) up to 3 times to explain the study and gauge interest in participation. Eligible and interested participants were scheduled for a videoconference meeting and provided informed consent on the day of their interview.

### Data collection and analysis

The interview guides were developed using a team-based approach and were revised iteratively during the initial interviews. We used similar interview guides for patients and clinicians (See Supplemental Materials, S1) that asked participants about their experience using the tool. We kept the interview guide questions broad and open-ended, but they were structured to probe for key tenets of the “people-like-me” approach (e.g., personalized care, patient-clinician communication, shared decision making).^13^

Interviews were conducted jointly by study team members (LC and JG) using a videoconferencing platform (Zoom® for patients and Microsoft Teams version 1.6.00.7354, for clinicians). After each interview, a member of the interview team completed a debriefing form (see Supplemental Materials, S2) to summarize emergent concepts, and patterns and to facilitate an iterative approach to revising the interview guide. Interviews were audio-recorded and transcribed verbatim, and transcripts were checked against the audio recordings for accuracy by a study team member.

Two members of the research team (LC and JG) analyzed the data immediately after each interview under the guidance of a senior qualitative researcher (MM). We analyzed patient and clinician interviews separately using descriptive content analysis that combined inductive and deductive approaches.^19,20^ We used Dedoose software (version 9.0.90) for all coding and analysis procedures.

First, we created a preliminary coding framework using the first four patient interviews. This involved a collaborative process among the analysts (JG and LC) where we reviewed the transcripts together, identified relevant sections of the text, and created preliminary codes. These preliminary codes were all derived from the data. However, given the structure of the interview guides, some of these codes encompassed key aspects of the “people-like-me” approach. We sought feedback from an experienced qualitative researcher (MM) to check our interpretation and labelling of codes to specific meaning units in the text. After we completed this process with the first four interviews, we refined our coding framework where we grouped key related codes into overarching themes and subsequently defining each code and theme. We then reviewed our coding framework and definitions with MM. Once we had a robust and well-defined coding framework, LC and JG independently coded these first 4 interviews again applying the coding structure, reviewing codes for consistency, and resolving any discrepancies of interpretation by consensus.

After establishing sufficient inter-coder reliability^21^, either LC or JG independently coded the remaining interviews. LC, JG, and MM met throughout the analysis process to discuss emerging patterns, reorganize the codebook, and identify any additional themes. Finally, we reviewed and refined our themes to ensure coded extracts reflected a coherent pattern and re-read the entire dataset to ensure final themes were reflective of the data. We used a similar approach for the clinician interviews. We interviewed patients until we reached thematic saturation^22^, and we interviewed all clinicians who expressed interest in participating.

## Results

Between October 2021 and May 2023, 16 patients and 10 clinicians were interviewed. Further information regarding recruitment, screening, and participant characteristics are available in Supplemental Materials (S3, S4). We identified four common main themes among patients and clinicians; therefore, we present results from these groups together. The common main themes cover: 1) Expectations, 2) Motivation, 3) Influence on practice and 4) Clarity and comprehension. Within these themes, subthemes were explored further (Table 1). In some cases, subthemes similarly and equally represent patient and clinician data, in other cases the subthemes are most representative of either a patient or clinician perspective, which we highlight narratively throughout.

**Table 1 -.** Themes and subtheme definitions.

| Theme 1: Motivation |  |
| --- | --- |
| Subtheme | Definition |
| *Positive reinforcement* | Positive reinforcement is a concept that encourages or rewards desirable behaviour and outcomes and offers a form of extrinsic motivation. |
| *Competition* | Competition is the act or process of trying to obtain a higher level of success than oneself or others, which can serve to increase motivation. |
| Theme 2: Expectations |  |
| Subtheme | Definition |
| *Setting expectations* | Setting expectations refers to the perception that the tool helped patients understand the projected course of their recovery. |
| *Meeting expectations* | Meeting expectations refers to the perception that the tool helped patients understand if their recovery was progressing satisfactorily relative to their peers. |
| Theme 3: Influence on practice |  |
| Subtheme | Definition |
| *Guiding treatment* | Guiding treatment describes how the tool helped to inform or direct patient treatment. |
| *Education tool* | Education tool refers to how the tool was used as an informational aid to reinforce key aspects of rehabilitation with patients. |
| **Theme 4: Clarity and comprehension** |  |
| **Subtheme** | **Definition** |
| *Understanding the “take-home” message* | Understanding the “take-home” message describes patient and clinicians’ ability to comprehend the general concept or main information provided by the tool. |
| *Health literacy* | Health literacy describes patients’ ability to understand the reference charts and was impacted by their baseline ability to interpret health information, graphs, and numbers. |

### 1) Expectations

Expectations are beliefs about anticipated outcomes. This includes how the tool helps inform whether patients are meeting projected benchmarks and how the tool shapes patients’ understanding of their projected course of recovery. Within the ‘Expectations’ theme we identified 2 subthemes (‘Meeting expectations’, and ‘Setting expectations’).

#### Meeting expectations

Patients consistently stated that the tool provided objective evidence that clarified if their recovery was progressing, on track, on par, or better than their peers. Patients noted that the tool’s feedback was particularly helpful given that the course of recovery is long, and it can be challenging to recognize progress.

“It’s just the reinforcement that, that it’s [rehabilitation] working. That this isn’t all or nothing. That, as tough as it seems day-to-day, that you were getting where you wanted to be.” (Participant 3, Patient)

Clinicians also felt the tool helped patients understand if their recovery was on track and gave them an objective way to measure progress, which was particularly impactful when there was a mismatch between patient and clinician perceptions of recovery. In some cases, this allowed clinicians to engage patients in a more meaningful dialogue that combined their clinical judgement with impartial evidence from the tool. One clinician commented that this was especially important because patients often compare their recoveries to friends or family who underwent TKA, which may lead to unrealistic expectations. Similarly, many clinicians stated the tool helped to validate their recommendations and promote greater buy-in from patients regarding the treatment plan.

“Oh, you’re just telling me that, you tell everybody that, I’m like, ‘No! Look! There’s data. Look, you are really right on par with what we’re expecting, or you really are ahead of where we’re expecting.’” (Participant 1, Clinician)

Some clinicians also reported using the tool with patients who were not meeting expectations to highlight targets for improvement.

#### Setting expectations

Some clinicians stated they used the tool pre-operatively to help set expectations surrounding anticipated trajectories of recovery. For example, a few clinicians stated that the tool encouraged pre-operative conversations regarding what to expect for post-operative ROM recovery.

“So, I think the really positive ones were really encouraging and you could kind of see people visibly like relax a little bit about it. Um, and then I think on the same, but opposite side, it gave realistic expectations to the people whose numbers were not good.” (Participant 3, Clinician)

Further, one clinician discussed using the tool to help patients revise their expectations surrounding post-operative knee pain.

“I know you’re still in pain. But look, everyone’s still in pain. You know, like using that, to normalize what they’re feeling. Because once again, their expectations may have been off.” (Participant 1, Clinician)

Patients discussed using the tool to set expectations less frequently than clinicians. One patient explained that they had unrealistic expectations before surgery about the speed of their recovery. Although their actual recovery was slower than anticipated, the tool assured them they were progressing satisfactorily. Another patient who had a prior knee replacement highlighted how the tool could help address some of the anxiety related to not knowing what to expect after surgery.

“And especially with the first [knee replacement], you don’t know what’s expected. They can tell you what they want to tell you, but you’re just worried about getting your knee better. And if you know where you actually are, and if you can see, ‘Okay, these are 100 people and I fall here. I’m doing okay, or I need to work more on this.’ That helps.” (Participant 11, Patient)

### 2) Motivation

Motivation refers to the tool’s influence on patients’ willingness or desire to participate in rehabilitation after TKA. Within the ‘Motivation’ theme, we identified 2 subthemes (‘Positive reinforcement’, and ‘Competition’).

#### Positive reinforcement

Often, patients who felt motivated by the tool discussed the concept of positive reinforcement. For example, patients discussed that reviewing the tool’s output was like getting a “pat on the back”, or a “star on my chart”.

Many patients stated the tool motivated them to continue participating in rehabilitation as it provided evidence that they were making progress in their recovery. Patients often stated that when the tool indicated they were meeting or exceeding expectations, it provided a form of validation or encouraged them to strive for an even better outcome.

“It was a wow factor. You know, I hadn’t seen those before. And it just gave me a lot of insight as to this is where I am now, and I know what to look forward to. And yeah, it’s a poor substitute for a coach, kicking your butt to get moving, but it’s a good second one.” (Patient, Participant 6)

In contrast, one participant who had a particularly hard time achieving range-of-motion after surgery described feeling demotivated or discouraged by the tool.

“Well, I wanted to do better when I saw that I wasn’t doing well, in some of those areas, but I couldn’t. My leg wouldn’t do it. I guess, yeah, you’re bummed because it’s just like getting a poor grade in school. You know, you missed the mark.” (Patient, Participant 10)

Clinicians similarly perceived that certain subgroups of patients were either motivated or demotivated by the tool. For example, when the tool suggested a patient was not meeting expectations, clinicians felt this could either motivate them to improve their status or discourage them depending on the patient.

Two clinicians indicated that in rare cases, they chose not to share the tool’s output with patients who were struggling in their recovery. A few clinicians also suggested they would prefer to use their clinical judgement in the future to determine if/how to use the tool’s output with certain patients, to promote positive patient interactions.

“I think it’s just, it’s not helpful at that point. Like, ‘hey, we know you’re not doing well. We know this is the direction you’re headed.’ I don’t think they need another piece of paper to tell them that.” (Participant 4, Clinician)

#### Competition

Patients often described that the tool gave them a sense of “winning”, accomplishment, or success in their recovery.

“I liked to see where I was. So that it would encourage me to push harder to… to either improve or get higher on the graph. To win more.” (Participant 4, Patient)

Clinicians perceived that certain patients were more likely to be motivated by the tool. For example, someone with a “type-A personality” or someone who enjoys a target to work towards (i.e., achieving a high score), was thought to be more motivated by the tool.

“If they were like goal-oriented people who like, needed a number to kind of work towards…it was like, ‘OK, well, here’s your number that you need to work for, for flexion or extension, that’s what we’re shooting for.’” (Participant 2, Clinician)

### 3) Influence on practice

Influence on practice refers to perceptions of how the tool impacted patients’ care during their rehabilitation after TKA. Within the ‘Influence on practice’ theme we identified 2 subthemes (‘Guiding treatment’, and ‘Education tool’).

#### Guiding treatment

Patients had mixed perceptions of how the tool was used to guide their care. Some patients indicated the tool supported collaborative decision making with their clinician, including which exercises to focus on or when to consider discharge from physical therapy.

“So, we had decided, and the graph supported that decision, I think, to go ahead and graduate me.” (Participant 13, Patient)

Other patients did not perceive that the tool influenced their plan of care directly, or they were unsure if it impacted their PT’s decision making. A few patients stated that the tool was not influential or emphasized during their rehabilitation and expressed a lack of interest in the tool.

“I really didn’t use the chart. I looked at it two times in that whole timeframe. I mean, I don’t know how I would have felt about it. I don’t know. I didn’t use it. You know, you just said, “You’re here, they’re there and you know, you’re done with it.” (Participant 10, Patient)

Clinicians similarly had mixed perceptions regarding the tool’s usefulness for guiding treatment. One clinician mentioned that the tool facilitated consistency with outcome assessments such as the TUG. Some clinicians felt the tool was helpful for guiding decisions regarding the frequency of care and for discussing these decisions with patients.

“So, if somebody was projected to be behind, it was information we then used at the eval to see if they still were, right? And that’s a higher frequency. That’s a ‘three time-a-week-er’, for sure, because we’re behind on range of motion, we need the reps, we need the eyes.” (Participant 3, Clinician)

However, most clinicians did not feel the tool influenced their treatment plan because it only validated what they had already planned, they used it inconsistently, or because they did not need it given their clinical experience treating this population. These clinicians often mentioned that the tool could be helpful to clinicians with less experience. This idea was reinforced by a clinician who stated the tool supported their clinical communication and helped to shape their practice in an early phase of their career.

“Probably given the timing of my career, it was also influencing for me to gain confidence in my communication skills. Like, I think it really impacted my ability to deliver the message of, “yes, you’re doing well. This is what we need. No, you’re not doing well. This is what we need or you’re right where we want you. And we’re confident in X amount of weeks you’re going to get there.” (Participant 6, Clinician)

#### Education tool

Clinicians emphasized that the tool enabled more consistent delivery of patient education and helped them prioritize discussions with patients about recovery and treatment.

“The patient obviously is gonna get some really good information during that time. But from the clinician aspect, it really forces the clinician to kind of take a second, step back, and, and be involved in that education process.” (Participant 7, Clinician)

Several clinicians stated that the tool helped to create educational opportunities and emphasize important elements of rehabilitation. Clinicians reported using the tool to validate their clinical judgement or to reinforce and consolidate patient education.

“I keep going back to like the education, but it backs up all the things we’ve always said. But now, it’s more believable because there’s a paper that shows it.” (Participant 3, Clinician)

### 4) Clarity and comprehension

Clarity and comprehension are concepts related to patient and clinician understanding and ability to interpret information provided by the tool. Within the ‘Clarity and comprehension’ theme we identified 2 subthemes (‘Understanding the “take-home” message’, and ‘Health literacy’).

#### Understanding the “take-home” message

Most patients reported a basic understanding of the tool’s purpose, and felt they were able to interpret the reference charts during their rehabilitation. Patients frequently commented that they understood if they were meeting expectations based on the chart.

“No, I think it was fairly self-evident. Lots of people, may have a problem in understanding with what 90 percentile means, or 10 percentile means. But even without that, the lines that are drawn, I think, if you’re in the middle, it’s almost, it’s intuitive, you know, that you’re hanging in with everybody else.” (Participant 6, Patient)

Some patients highlighted factors that supported their understanding of the tool, such as having a data intense occupation or reviewing the charts with their physical therapist.

Patients also appeared to have a basic understanding that the tool was comparing them to similar people, however, the details surrounding these comparisons were not always clear. Patients frequently recalled they were being compared to people who had the same surgery and were a similar age, but few mentioned other matching variables used by the tool (e.g., sex, body mass index, preoperative status).

Overall, clinicians felt that the reference charts generated by the tool were easy to understand and use with patients, but they also identified areas for improvement to make the charts more accessible and interpretable for patients (described in next section).

#### Health literacy

Health literacy describes patients’ ability to understand the information provided by the tool and was impacted by their baseline ability to interpret health information, graphs, and numbers. Many patients reported being confused about aspects of the tool including its use of percentiles, line graphs, and medical jargon. Other patients could not recall what outcome measures were tracked by the tool.

“It’s just like, when you see a graph paper, it’s just checks. And it doesn’t mean anything to you, unless you write something on it. Especially old people. You have to draw a picture.” (Participant 12, Patient)

Patients offered suggestions to improve the interpretability of the charts including simplifying the “take-home” message, using more words to explain or accompany the reference charts, avoiding acronyms such as TUG, and the use of color.

Clinicians also reported that some patients were unable to meaningfully engage with or interpret the charts depending on their health literacy or prior experience with physical therapy.

“But I do think If they don’t have that underlying knowledge, some of the meaning of it might not be as relevant to them if they don’t quite get what the curve is… I don’t feel like they necessarily understood where they were, they would take my word for it, but they couldn’t look at the graph and be like, to be able to interpret that.” (Participant, Clinician 1)

Clinicians commented that some patients had difficulty interpreting what was considered a positive or negative trajectory on the chart depending on the outcome measure. For example, a lower score on the TUG and a higher degree of knee flexion both represent positive improvement. Additionally, clinicians explained that the reference charts had to be printed in black and white at their clinics, which reduced the interpretability of the color-coded legend. Overall, these challenges were perceived to decrease patients’ clarity, comprehension, and engagement with the tool. Clinicians provided suggestions to make the charts more patient friendly including adding pictures of a knee to increase the interpretability of joint angles, indicating what is a positive or negative trend on the reference chart using pictures, and linking outcome measures such as the TUG to functional tasks (e.g., walking or stairs).

## Discussion

Overall, this study identified four themes related to patient and clinician experiences with the “people-like-me” tool including: 1) Expectations, 2) Motivation, 3) Influence on practice and 4) Clarity and comprehension.

Most patients and clinicians felt that the tool provided patients with valuable feedback about their recovery progress and whether they were meeting expectations for recovery. Clinicians also highlighted that they used the tool to set patient expectations about their recovery. These findings parallel the results of Sinnige et al.’s recent vignette study that examined a similar approach to “people-like-me”. Sinnige et al. explored PT’s perceptions of using personalized outcomes forecasts for patients with intermittent claudication.^15^ Similar to our study, they found that PTs thought personalized recovery predictions could be useful for setting patient expectations in rehabilitation.^15^ These findings are particularly meaningful for post-TKA care, because unmet expectations are a leading cause of patient dissatisfaction,^2,23,24^ and patients’ preoperative expectations tend to be overly optimistic.^25^

Interestingly, we found that mismatches between patient and clinician perceptions of recovery could be addressed by the tool. Clinicians felt it helped them to communicate more effectively about patient recovery by combining their clinical judgement with impartial evidence. This allowed patients to visualize and understand their individual prognosis and progress over the course of rehabilitation which, in some cases, helped alleviate anxiety related to whether they were “on track” during their recovery. In this way, the tool may help patients renegotiate or revise their expectations during the post-operative period, which could improve patient satisfaction after TKA. In contrast, clinicians traditionally use generic benchmarks (e.g., postoperative protocols) to communicate with patients about their recovery, which may be less applicable or interpretable for some patients.

Helping patients understand their personalized recovery trajectory may be especially important given that patients frequently draw on the experiences of family and friends when conceptualizing their expectations of TKA recovery^26,27^—even though they recognize this is not necessarily a valid approach.^27^ In a qualitative investigation, Goldsmith et al. identified that individuals who have undergone TKA want information on a variety of recovery trajectories “so they could be assured they were on some sort of a track to recovery, even if it was not the ideal track or an ideal recovery”, and because this information is not readily available, they often rely on comparing themselves to friends and family who have undergone TKA.^26^ Goldsmith et al. participants also expressed that it was challenging when their recovery experience was worse than others and that they wanted support from clinicians in understanding this mismatch.^26^ Our study demonstrates that a tool that provides objective and personalized data for TKA recovery can help clinicians to address this informational need.

We also found that the tool may motivate patients to participate in rehabilitation by providing positive reinforcement and/or a form of competition. Relatedly, a recent qualitative study highlighted that positive reinforcement may increase motivation and exercise participation after TKA, as it provides feedback that can lead to improved self-regulation and goal attainment.^28^ Motivating patients in rehabilitation after TKA can be challenging because progress is gradual, variable, and often painful.^6–8^ Positive reinforcement via the tool may increase patient motivation to participate in rehabilitation and support patients to meet their personal goals after surgery.

While the tool’s motivational impact was mostly discussed as helpful, some patients and clinicians felt that the tool could be demotivating if it reminds struggling patients that they are not meeting expectations. In some of these cases, clinicians stated they may opt not to use the tool with patients. Similarly, in Sinnige et al.’s study, therapists perceived that providing patients with personalized recovery information could motivate patients to achieve therapy goals or could demotivate patients with a poor prognosis.^15^ Some therapists from their study indicated they wouldn’t provide personalized recovery information to individuals who were performing below predicted measurements.^15^ Sinnige et al. discussed that withholding information from patients because of a poor prognosis may be problematic given patients’ right to know about their health, yet they also acknowledged that the use of clinical tools is often at the discretion of clinicians.^15^ We theorize the motivational impact of the tool likely depends on individual patient characteristics (i.e., goal-oriented) and the unique details of the patient’s recovery experience. If clinicians are concerned that a patient will react negatively to the tool, it may be prudent to simply ask the patient if they want that information.

Participants’ perceptions of the tool’s influence on practice varied. Some patients and clinicians indicated that it helped guide decisions related to exercise selection, frequency of care, and discharge. Many clinicians also stated the tool created consistent opportunities to educate patients on their recovery status and facilitated discussions surrounding their care. These results suggest the tool has the potential to support a collaborative and personalized approach to rehabilitation. However, most patients and clinicians did not perceive that the tool was used to directly inform treatment decisions or to support shared decision making. Sinnige et al. similarly found that PTs did not envision using personalized outcomes forecasts to modify treatments or to support shared decision making.^29^ These results suggest that intentional training approaches are needed to promote these types of clinician behaviors. In the current study, we used self-paced online training modules to orient clinicians to the “people-like-me” tool and demonstrate how it could support clinical decisions and shared decision making.^18^ However, recent evidence suggests that active training approaches that incorporate performance feedback and/or mentored interactions are more effective for changing physical therapists’ behaviors.^30,31^ Further, our training did not incorporate a formal shared decision making framework or theory. In a follow up to their vignette study, Sinnige et al. demonstrated that providing physical therapists with in-depth, theory-driven training for using personalized outcomes forecasts can increase shared decision making.^32,33^

Finally, the use of percentiles, line graphs, and medical jargon were thought to decrease patients’ clarity and comprehension of the tool. Relatedly, in their national survey, Ben-Joseph et al. demonstrated that many U.S adults do not understand how to interpret pediatric growth charts,^34^ which closely resemble the reference charts created by the tool. In future iterations, we plan to simplify the tool’s graphical and numerical output, use lay language more consistently (e.g., knee bending instead of flexion), and simplify the presentation of percentiles.

## Limitations

Our study has some limitations. Firstly, patients and clinicians who volunteered to participate may have held more favorable views of the tool. However, we feel that we were able to capture a balanced view of the tool including both positive/negative perceptions and areas for improvement. Further, we may find different perceptions of the tool based on regional or institutional differences, thus the transferability of these results to other settings is unknown. Our finding that most clinicians did not use the tool to guide treatment decisions or facilitate shared decision making may have been influenced by the training strategies used in this pilot study. Future investigations of this tool should consider using more intentional, theory-driven training approaches to promote these types of changes in clinician behavior. Relatedly, despite the mostly positive perceptions held by participants in this study, more research is needed to determine (a) whether this tool can be adapted to accommodate patients with lower levels of health literacy and (b) whether it can meaningfully impact clinical practice.

## Conclusions

We explored patients’ and clinicians’ perceptions of a clinical decision support tool that promotes personalized rehabilitation after TKA. Most patients and clinicians reported that the tool helped to shape patients’ expectations for postoperative recovery and increase patient motivation to participate in rehabilitation. Patients and clinicians had mixed perceptions on how the tool influenced clinical practice. Finally, most patients understood the tool’s “take-home” message, however, patients and clinicians identified some limitations in patient comprehension of the tool, which has prompted revisions to the tool to accommodate varying levels of health literacy.

## Supporting information

Supplemental Materials_Interview Guides

Supplemental Materials Participant Characteristics

Supplemental Materials_Interview Debriefing

Supplemental Materials_Flow diagram

## Data Availability

All data produced in the present study are available upon reasonable request to the authors

## Abbreviations

TKA: total knee arthroplasty
PT: physical therapist
ROM: range of motion
TUG: Timed Up and Go

## Acknowledgements

The authors would like to thank the following individuals for their contributions throughout this project: Adam D. Lutz, PT PhD, Dawn Waugh, PT, DPT, and Kathryn Van Sciver-Richardson

