## Supplemental Materials_Interview Guides for "Patient and Clinician Perceptions of a “People-Like-Me” Tool for Personalized Rehabilitation after Total Knee Arthroplasty: A Qualitative Interview Study"

Supplemental Materials (S1)- Interview Guides for Patients and Clinicians

**Interview Guide – Patients**

1. Please tell me about your experience in rehabilitation after total knee replacement.
    a. Probe: What was the most challenging aspect of your recovery?

2. Please tell me about your experience using the “people-like-me” tool

3. How were you introduced to the tool?

4. When you were initially informed about the tool, how did you feel about using it for your rehabilitation?
5. If you were to have another knee replacement, would you be interested in using the tool again? Why or why not?
6. Did the tool influence your recovery in any way?
    **a.** Probes
        **i.** Mobility
        **ii.** Pain
        **iii.** Range of motion
        **iv.** Positive and negative examples
        **v.** If outcomes were improved, what do you feel lead to this improvement?
7. What did you think about the charts generated by the tool?
    **a.** Probes
        **i.** Were the outcomes tracked by the app useful? Why or why not?
        **ii.** In a perfect world what could a chart tell you that would make you feel more informed or confident about your recovery after knee replacement surgery?
        **iii.** How important was it for you to be within the “normal” range on the charts?
8. How did using the tool impact your overall experience in rehab after TKA?
    **a.** Probes
        **i.** Hypothetical patient doing well <u>or</u> poorly
        **ii.** Anxiety / Expectations
        **iii.** Engagement
        **iv.** Positive and negative examples

**v.** Visit utilization (frequency/duration of care)

vi. If your experience was improved, what do you feel lead to this improvement?

9. How did your physical therapist use the tool with you during your rehabilitation?
   **a.** Probes
      **i.** Did using the tool ever change the plan for your treatment? How so?
      **ii.** How could have it been used more effectively?
      **iii.** When / how often did your physical therapist use it with you?

10. Was anything about the tool difficult for you to understand?
    **a.** Probes
       **i.** How could it have been improved?
       **ii.** Can you explain to me who you were being compared to on the charts?
          1. Probe: If “people-like-me” is unclear, tell patient they *were* being compared to similar patients. Does knowing this change the way you think about the charts? Would have changed your experience?
          2. Probe: Give example of how “people-like-me” are determined using age, sex, height/weight, preoperative status
11. Can you walk me through the details of one of your charts and explain to me how you felt about this information during your rehabilitation?

**GENERAL PROBES**

Can you explain in more detail?

Can you give us an example?

**Interview Guide – Providers**

1. What are some of the most challenging aspects of supporting patients recovering from knee replacement?
   a. How did you anticipate the “people-like-me” tool might help address some of these challenges?

i. Probe: To what extent did the tool meet your expectations?

2. How were you first introduced to the tool?
    a. Were you interested in using the tool? Why or why not?
3. What were patients' reasons for not wanting to use the tool?
    a. Probe: How could these reasons be addressed in the future?

4. What were some reasons that patients DID want to use the tool?
    a. How could these reasons be enhanced to encourage more patient participation?

5. How did using the tool influence the way you practiced for patients with TKA?
    a. Probe
        i. Utilization (frequency of care, discharge decisions)
        ii. Treatment priorities

6. How did using the tool influence outcomes for your patients?

7. How did using the tool influence patients' experience in rehab?
    a. Probes
        i. Anxiety
        ii. Engagement
        iii. Positive and negative examples
        iv. If patient experience was improved, what mechanisms lead to this improvement?

8. If up to you, would you continue using the tool after the study? Why or why not?

**GENERAL PROBES**

Can you explain in more detail?

Can you give us an example?
