## Supplemental Materials Participant Characteristics for "Patient and Clinician Perceptions of a “People-Like-Me” Tool for Personalized Rehabilitation after Total Knee Arthroplasty: A Qualitative Interview Study"

Supplemental Materials (S3)- Characteristics of participants and potential patient participants

| **Characteristic** | | |
|---|---|---|
| **Patients** | Patients who participated in interviews (n = 16) | Patients exposed to the tool (n = 167) |
| Age (years) | Median = 72<br>Range = 51, 84 | Median = 70<br>Range = 44, 89 |
| Sex distribution | Female = 8<br>Male = 8 | Female = 89<br>Male = 78 |
| **Clinicians who participated in interviews (n = 10)** | | |
| Clinical experience (years) | Median = 6.25<br>Range = 3.5, 21 | |
| Sex distribution | Female = 8<br>Male = 2 | |
