## Supplemental Materials_Interview Debriefing for "Patient and Clinician Perceptions of a “People-Like-Me” Tool for Personalized Rehabilitation after Total Knee Arthroplasty: A Qualitative Interview Study"

Supplemental Materials (S2)- Interview debriefing form[1]

Key informant interview:

Date interview conducted:

Today's date:

1. What were the main issues or themes that struck you during this interview?

2. What information seemed consistent with what you have learned from other interviews or data sources?

3. What information seemed to contradict what you have learned from other interviews or data sources?

4. What new questions emerged for you during this interview that you might want to explore with other key informants/ data sources?

5. Did the interview seem to flow well? In what areas? How could this be improved?

6. What interview questions seemed to work well?

**7.** What interview questions did <u>not</u> seem to work well? How might these questions be revised?

**8.** What else is important to capture about this interview?

[1] This form was modeled after Figure 4.1 *in* Miles, Matthew B. and Huberman, A. Michael. (1994). Qualitative Data Analysis, 2nd edition. Sage Publications, Thousand Oaks, CA. pp. 53.
