## Supplemental Materials_Flow diagram for "Patient and Clinician Perceptions of a “People-Like-Me” Tool for Personalized Rehabilitation after Total Knee Arthroplasty: A Qualitative Interview Study"

Supplemental Materials (S4)- Flow diagram demonstrating the number of participants contacted, screened, and ultimately included in the study.

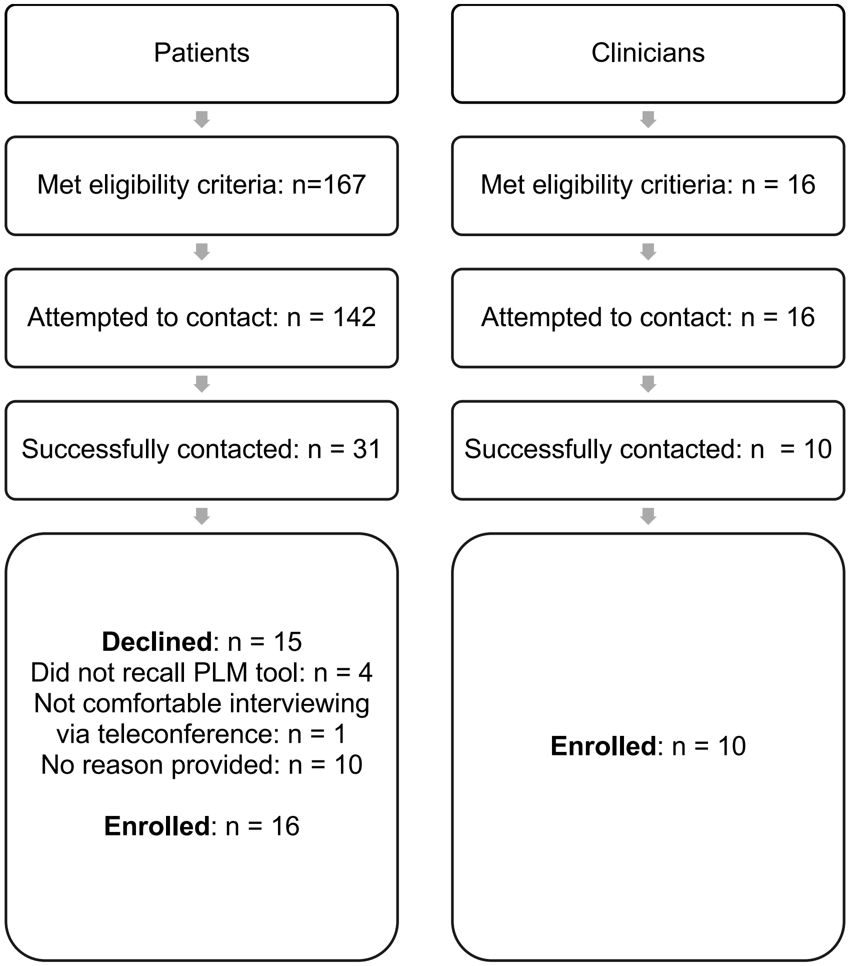
